# Analyzing the Spatial Distribution and Language Abilities of Physicians in Alberta, Canada

**DOI:** 10.1101/2023.09.14.23295573

**Authors:** Christopher Belanger, Lise M. Bjerre

## Abstract

This study combined public data and geospatial analysis to examine physicians’ language abilities and locations in Alberta, Canada, and produced an interactive map to allow patients and policymakers to view the data. We identified n=11,370 active physicians in the province of Alberta, of whom we further identified n=194 (1.7%) as University of Ottawa (uOttawa) graduates, n=955 (8.4%) as French-speaking, and n=4,965 (43.7%) as community-based family physicians. French-speaking physicians were concentrated in Census Division 6 (n=464, 48.6%) surrounding Calgary and Census Division 11 (n=356, 37.3%) surrounding Edmonton. Overall reported French-language ability was low, with just 955 (8.4%) of all active physicians reporting competency in French. uOttawa graduates (n=70, 36.1%) were much more likely to report French ability than graduates of other schools (n=885, 7.9%), women (n=457, 9.6%) were slightly more likely than men (n=497, 7.6%), and specialists (n=666, 10.4%) were more likely than family physicians (n=289, 5.8%).

## Introduction

The University of Ottawa’s Faculty of Medicine is interested in better understanding the distribution of French-speaking physician care in the province of Alberta, especially among uOttawa graduates. This study combined public data with geospatial analysis to learn how and where French-speaking uOttawa graduates practice in Alberta, and to produce an interactive online map to allow patients and policymakers to explore the data.

To better understand the supply and distribution of French-speaking physicians in Alberta, Canada, and especially of community-based family physicians, this study has four goals:

- Collecting data from the College of Physicians and Surgeons of Alberta;
- Analyzing the supply and distribution of family physicians;
- Analyzing the supply and distribution of French-speaking physicians; and,
- Producing an interactive map of Alberta family physicians searchable by language, analogous to prior work—see www.docmapper.ca / www.trouvezunmedecin.ca.

## Methods

### Data Collection

Data was collected from the official “Physician Search” website of the Alberta College of Physicians and Surgeons (CPSA) during the week of January 31, 2023 [3]. A search was run with fully default parameters, which returned 35,188 basic results with general information (name, location, practice discipline, gender, languages spoken, and web link for more details). For each basic result, we then followed the web link and collected more detailed information (phone number, fax number, specialties, membership status, conditions on practice, registration number, qualifications, and whether they reported accepting new patients). Finally, we collected latitude and longitude coordinates for each physicians’ location using Google’s geocoding API [4]. Initial geocoding failed for n=195 physicians whose addresses included PO boxes, so we removed PO boxes from their locations and geocoded the remaining address.

The final result was a tabular data set with 35,1888 rows and the following 15 columns: name, location, phone_number, fax_number, practice_disciplines, specialties, gender, languages, membership_status, conditions_on_practice_permit, registration_number, accepting_new_patients, qualifications, lat, lon.

Provincial boundaries were collected from Statistics Canada’s digital boundary files [5].

### Data Cleaning and Processing

We cleaned the data in several steps outlined in Figure 1. First, we removed all physicians whose membership status was not “Active.” There were ten other possible membership statuses in addition to “Active,” including “Inactive,” “Inactive - Deceased,” “Suspended,” and “Active - Not in current practice.” Removing n=19,933 physicians with these other membership statuses resulted in a set of 15,255 physicians with active registrations.

**Figure 1:**
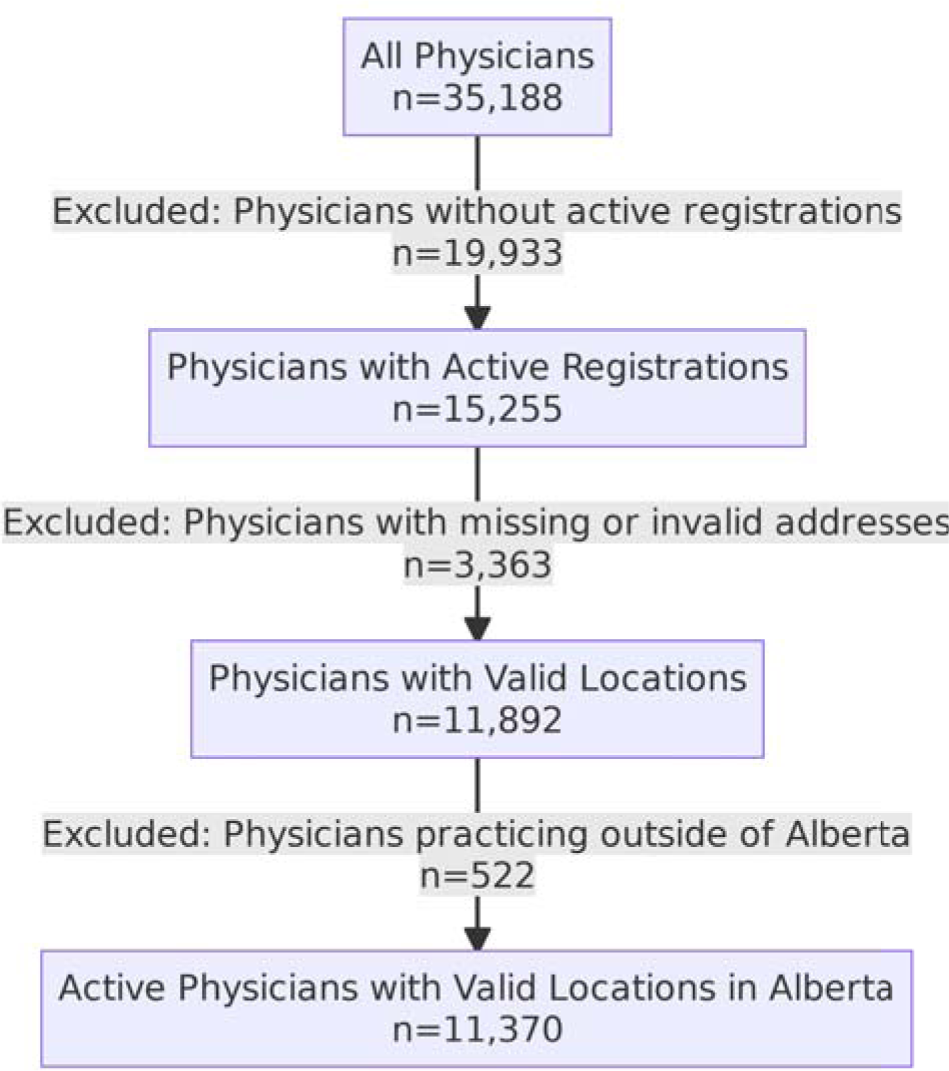
Population creation diagram for Alberta physicians dataset.

Next, we excluded n=3,363 physicians with missing or invalid addresses. This included n=3,333 physicians whose contact information had been intentionally withheld, listed only as “Contact information for this physician has not been made public.” This yielded a set of 11,892 active physicians with valid addresses.

Next we excluded n=521 physicians practicing outside the province of Alberta, using physicians’ geocoded latitudes and longitudes and keeping only those inside Statistics Canada’s digital boundary for the province of Alberta. We verified visually that the physicians excluded were all outside the province’s borders, and the physicians included were all inside. This yielded a final set of 11,370 active physicians with valid addresses in Alberta.

### Identifying Family Physicians

Beginning with the full set of n=11,370 active physicians, we further refined this set to identify active community-based family physicians. First, we excluded n=5,919 physicians whose listed specialties or practice disciplines did not include the terms “Family Medicine” or “General Practice”, leaving n=5,451 potential family physicians. We then excluded n=454 family physicians whose work location included the terms “hospital” or “emergency,” leaving 4,997 non-hospital-or-emergency-department-based potential family physicians. Before exclusion, we verified that there were no false positives, and that one ambiguous location, the Northern Lights Health Centre at 7 Hospital St. in Fort McMurray, does not list primary care services to the public on its website [6].

Our final set of n=4,997 family physicians indicates that 47.9% of Alberta’s registered physicians are providing community-based primary care services.

### Identifying University of Ottawa Graduates

Physicians who obtained their medical degree (MD) from the University of Ottawa were identified by excluding n=11,176 physicians without the word “Ottawa” in their degree information, leaving n=194 University of Ottawa Graduates as shown in Figure 3.

**Figure 2:**
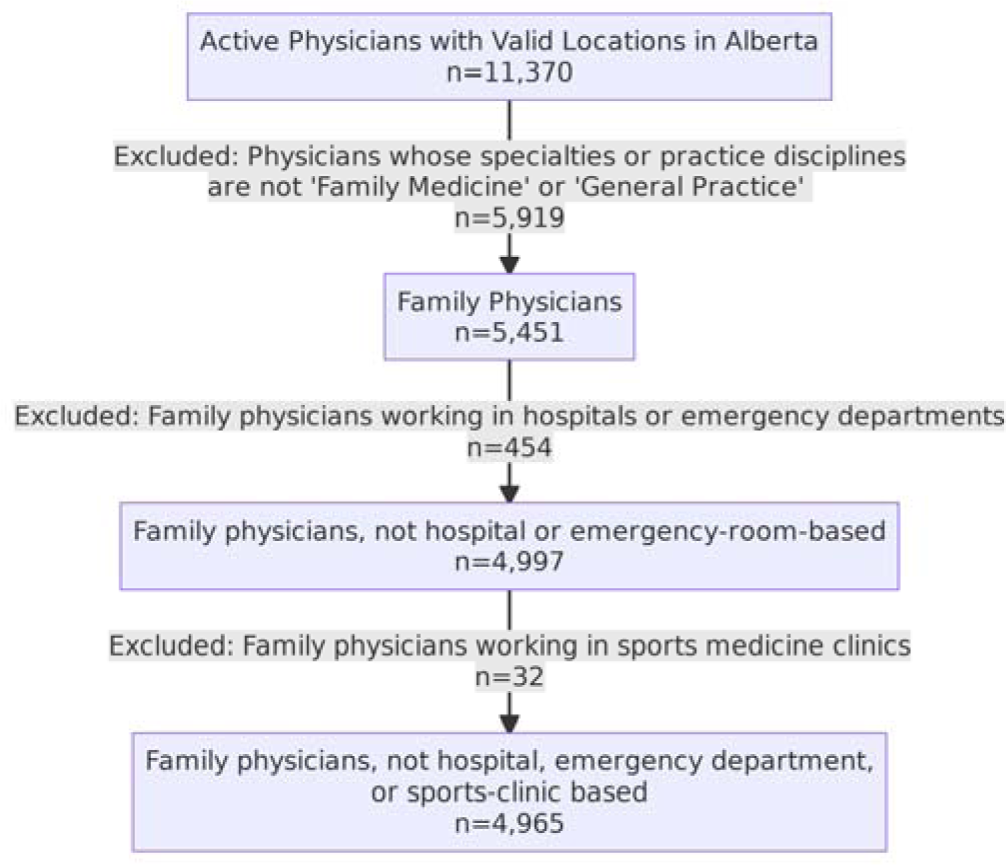
Population creation diagram for Alberta family physicians dataset.

**Figure 3:**
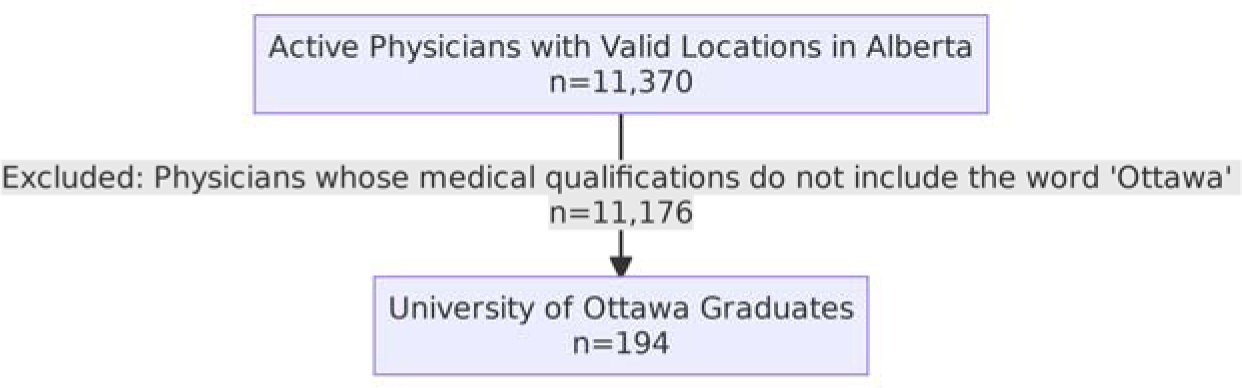
Population creation diagram for University of Ottawa graduates data set.

Note that this only includes physicians who received their medical degree from the University of Ottawa, and so excludes, for example, those who completed their residency requirements at the University of Ottawa. Residency and other qualification information is not provided by the CPSA. Identifying Alberta-based physicians who completed residencies or other qualifications at the uOttawa would require internal uOttawa data, for example physician names and specialties, and additional analysis to link the CPSA data with the uOttawa data.

### Software Used

All analysis described in this report was done using the R Language for Statistical Computing [1] and RStudio [2]. These are open-source software packages that enable code-based and reproducible research: since all steps in the analysis are executed through computer code, anyone with access to the code and input data is able to inspect all analytical steps and to reproduce them for themselves. The code used in this analysis can be made available upon request.

## Results

In this section, we present summary tables and visualizations describing our main results. We begin with population characteristics for the overall physician cohort, followed by more detailed tables for several subcategories of interest (family physicians, French-speaking physicians, and uOttawa graduates). We also provide more focused tables exploring French-speaking ability and physician locations.

### Population Characteristics

#### All Physicians

Abbreviated population characteristics for the full set of active physicians are shown in Table 1. Categories with more than five values have been truncated to the top five.

**Table 1:** Population characteristics for all n=11,370 physicians.

| Characteristic (Total n=11370) | n | % |
| --- | --- | --- |
| **Gender** |  |  |
| Female | 4749 | 41.8% |
| Male | 6575 | 57.8% |
| Unknown | 46 | 0.4% |
| **Language Spoken\* (n=108 distinct responses)** |  |  |
| English | 11370 | 100.0% |
| French | 955 | 8.4% |
| Afrikaans | 612 | 5.4% |
| Arabic | 604 | 5.3% |
| Hindi | 597 | 5.3% |
| Other | 3849 | 33.9% |
| **Practice Discipline\*\* (n=109 distinct responses)** |  |  |
| Family Medicine | 3864 | 34.0% |
| General Practice | 1259 | 11.1% |
| Internal Medicine | 1144 | 10.1% |
| Psychiatry | 644 | 5.7% |
| Pediatrics | 594 | 5.2% |
| Other | 3865 | 34.0% |
| **Institution / Country of Degree\*\* (n=178 distinct responses)** |  |  |
| UofA - University of Alberta | 2787 | 24.5% |
| UofC - University of Calgary | 1694 | 14.9% |
| South Africa | 747 | 6.6% |
| University of British Columbia | 485 | 4.3% |
| University of Saskatchewan | 442 | 3.9% |
| Other | 5215 | 45.9% |

\* Respondents could report competency in more than one language.
\*\* Values are verbatim responses.

#### Family Physicians

Population characteristics for the set of family physicians are presented in Table 2. Categories with more than five values have been truncated to the top five.

**Table 2:** Population characteristics for n=4,965 community-based family physicians.

| Characteristic (Total n=4965) | n | % |
| --- | --- | --- |
| **Gender** |  |  |
| Female | 2325 | 46.8% |
| Male | 2594 | 52.2% |
| Unknown | 46 | 0.9% |
| **Language Spoken\* (n=98 distinct responses)** |  |  |
| English | 4965 | 100.0% |
| Afrikaans | 503 | 10.1% |
| Arabic | 354 | 7.1% |
| Hindi | 329 | 6.6% |
| Punjabi | 298 | 6.0% |
| Other | 1897 | 38.2% |
| **Practice Discipline\*\* (n=12 distinct responses)** |  |  |
| Family Medicine | 3579 | 72.1% |
| General Practice | 1227 | 24.7% |
| Family Medicine (Emergency Medicine) | 87 | 1.8% |
| Family Medicine (Palliative Care) | 16 | 0.3% |
| Family Medicine (Family Practice Anesthesia) | 13 | 0.3% |
| Other | 43 | 0.9% |
| **Institution / Country of Degree\*\* (n=137 distinct responses)** |  |  |
| UofA - University of Alberta | 1218 | 24.5% |
| UofC - University of Calgary | 685 | 13.8% |
| South Africa | 601 | 12.1% |
| Nigeria | 292 | 5.9% |
| Libya | 191 | 3.8% |
| Other | 1978 | 39.8% |

\* Respondents could report competency in more than one language.
\*\* Responses were tabulated as given and may contain inconsistencies.

#### French-Speaking Physicians

Abbreviated population characteristics for n=955 French-speaking physicians are shown below in Table 3. Categories with more than five values have been truncated to the top five.

**Table 3:**
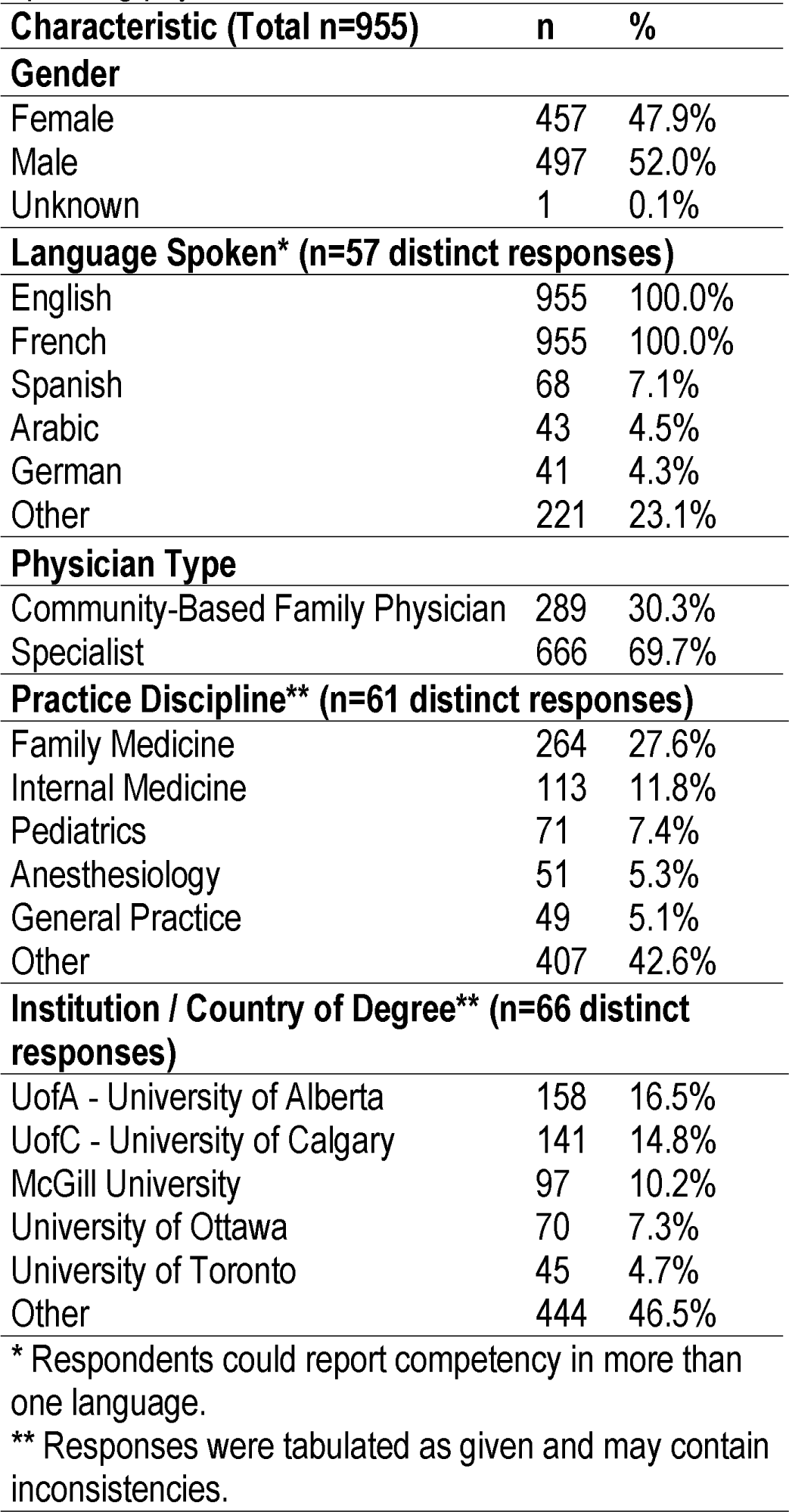
Population characteristics for n=955 Frenchspeaking physicians.

| Characteristic (Total n=955) | n | % |
| --- | --- | --- |
| **Gender** |  |  |
| Female | 457 | 47.9% |
| Male | 497 | 52.0% |
| Unknown | 1 | 0.1% |
| **Language Spoken\* (n=57 distinct responses)** |  |  |
| English | 955 | 100.0% |
| French | 955 | 100.0% |
| Spanish | 68 | 7.1% |
| Arabic | 43 | 4.5% |
| German | 41 | 4.3% |
| Other | 221 | 23.1% |
| **Physician Type** |  |  |
| Community-Based Family Physician | 289 | 30.3% |
| Specialist | 666 | 69.7% |
| **Practice Discipline\*\* (n=61 distinct responses)** |  |  |
| Family Medicine | 264 | 27.6% |
| Internal Medicine | 113 | 11.8% |
| Pediatrics | 71 | 7.4% |
| Anesthesiology | 51 | 5.3% |
| General Practice | 49 | 5.1% |
| Other | 407 | 42.6% |
| **Institution / Country of Degree\*\* (n=66 distinct responses)** |  |  |
| UofA - University of Alberta | 158 | 16.5% |
| UofC - University of Calgary | 141 | 14.8% |
| McGill University | 97 | 10.2% |
| University of Ottawa | 70 | 7.3% |
| University of Toronto | 45 | 4.7% |
| Other | 444 | 46.5% |

\* Respondents could report competency in more than one language.
\*\* Responses were tabulated as given and may contain inconsistencies.

#### uOttawa Graduates

Abbreviated population characteristics for n=194 uOttawa graduates are shown in Table 4. Categories with more than five values have been truncated to the top five.

**Table 4:** Population characteristics for n=194 uOttawa graduates.

| Characteristic (Total n=194) | n | % |
| --- | --- | --- |
| **Gender** |  |  |
| Female | 100 | 51.5% |
| Male | 94 | 48.5% |
| **Language Spoken\* (n=25 distinct responses)** |  |  |
| English | 194 | 100.0% |
| French | 70 | 36.1% |
| Hindi | 6 | 3.1% |
| Polish | 6 | 3.1% |
| Mandarin | 4 | 2.1% |
| Other | 33 | 17.0% |
| **Physician Type** |  |  |
| Community-Based Family Physician | 53 | 27.3% |
| Specialist | 141 | 72.7% |
| **Practice Discipline\*\* (n=37 distinct responses)** |  |  |
| Family Medicine | 55 | 28.4% |
| Internal Medicine | 30 | 15.5% |
| Anesthesiology | 11 | 5.7% |
| Pediatrics | 11 | 5.7% |
| Diagnostic Radiology | 10 | 5.2% |
| Other | 77 | 39.7% |
| **Institution / Country of Degree\*\* (n=1 distinct responses)** |  |  |
| University of Ottawa | 194 | 100.0% |
| Other | 0 | 0.0% |

\* Respondents could report competency in more than one language.
\*\* Responses were tabulated as given and may contain inconsistencies.

### French-Language Ability

Table 5 below shows French-language ability for different physician categories. Overall, only 955 (8.4%) of all active physicians reported competency in French. However, there was substantial variation between groups, with family physicians less likely to report French ability (5.8%) and uOttawa graduates more likely (36.1%).

**Table 5:** French-speaking ability among physician subcategories.

| Characteristic (Total n=11370) | French-Language Ability | n | % |
| --- | --- | --- | --- |
| **Gender** |  |  |  |
| Female (n=4749) | French-Speaker | 457 | 9.6% |
|  | Non-French-Speaker | 4292 | 90.4% |
| Male (n=6575) | French-Speaker | 497 | 7.6% |
|  | Non-French-Speaker | 6078 | 92.4% |
| Unknown (n=46) | French-Speaker | 1 | 2.2% |
|  | Non-French-Speaker | 45 | 97.8% |
| **Physician Type** |  |  |  |
| Community-Based Family Physician (n=4965) | French-Speaker | 289 | 5.8% |
|  | Non-French-Speaker | 4676 | 94.2% |
| Specialist (n=6405) | French-Speaker | 666 | 10.4% |
|  | Non-French-Speaker | 5739 | 89.6% |
| **Degree-Granting Institution** |  |  |  |
| Other Institution (n=11176) | French-Speaker | 885 | 7.9% |
|  | Non-French-Speaker | 10291 | 92.1% |
| University of Ottawa (n=194) | French-Speaker | 70 | 36.1% |
|  | Non-French-Speaker | 124 | 63.9% |

### Location

As can be seen in Figures 4-7, the majority of physicians in all categories were located in Census Division 6 surrounding Calgary and Census Division 11 surrounding Edmonton. Further details are shown below in Table 6.

**Table 6:** Census division locations for physician subcategories.

| Physician Category | Calgary / Div. 6 | Edmonton / Div. 11 | Other |
| --- | --- | --- | --- |
| All | 5030 (44.2%) | 4364 (38.4%) | 1976 (17.4%) |
| Family Physicians | 2180 (43.9%) | 1643 (33.1%) | 1142 (23.0%) |
| French-Speaking | 464 (48.6%) | 356 (37.3%) | 135 (14.1%) |
| uOttawa Graduates | 111 (57.2%) | 59 (30.4%) | 24 (12.4%) |

**Figure 4:**
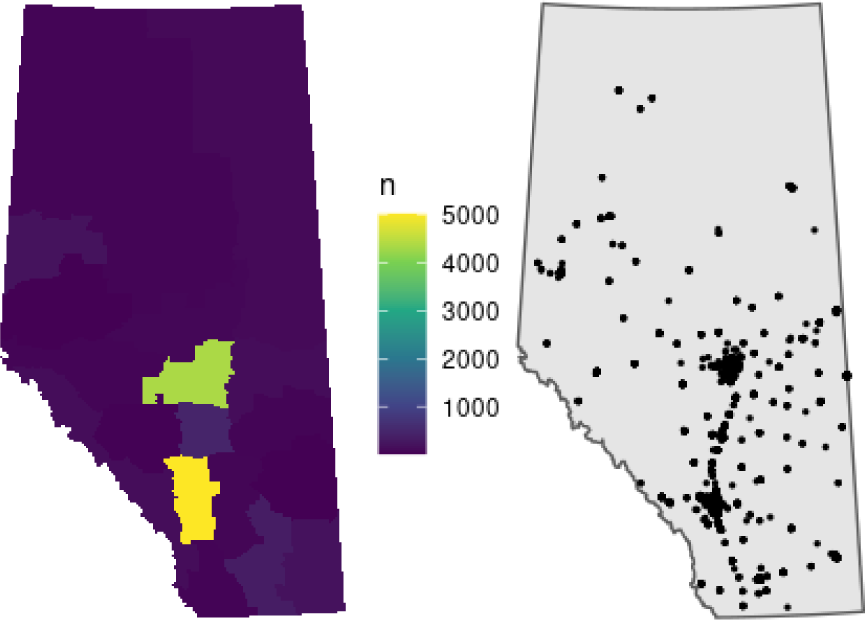
Distribution of all active physicians in Alberta, density by census division (left) and point distribution (right).

**Figure 5:**
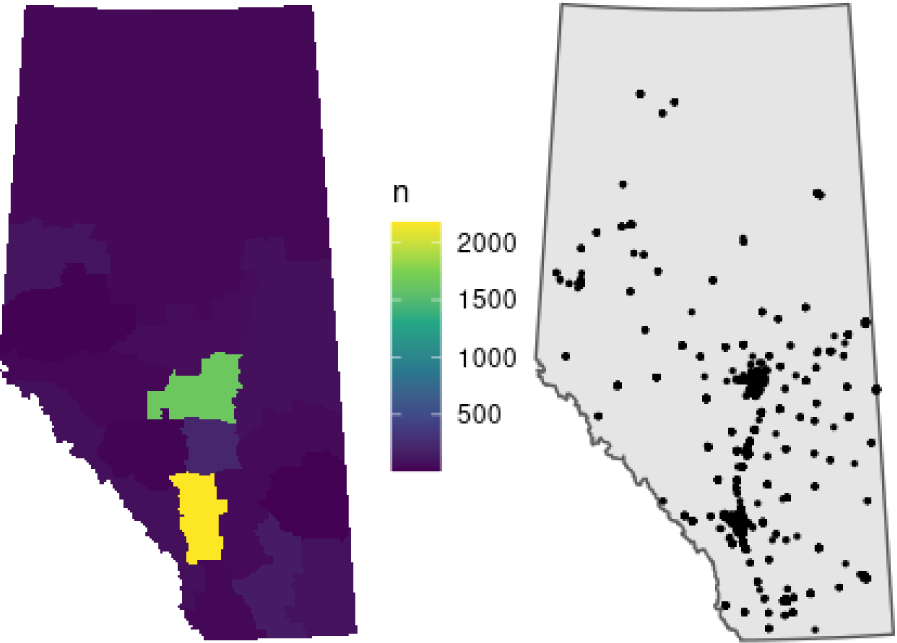
Distribution of active communitybased family physicians in Alberta, density by census division (left) and point distribution (right).

**Figure 6:**
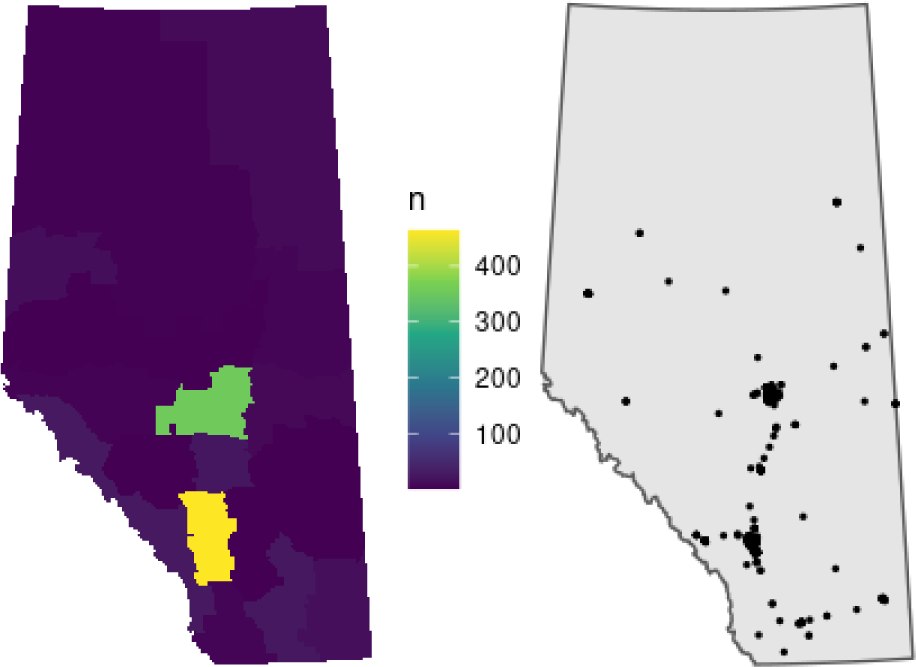
Distribution of active French-speaking physicians in Alberta, density by census division (left) and point distribution (right).

**Figure 7:**
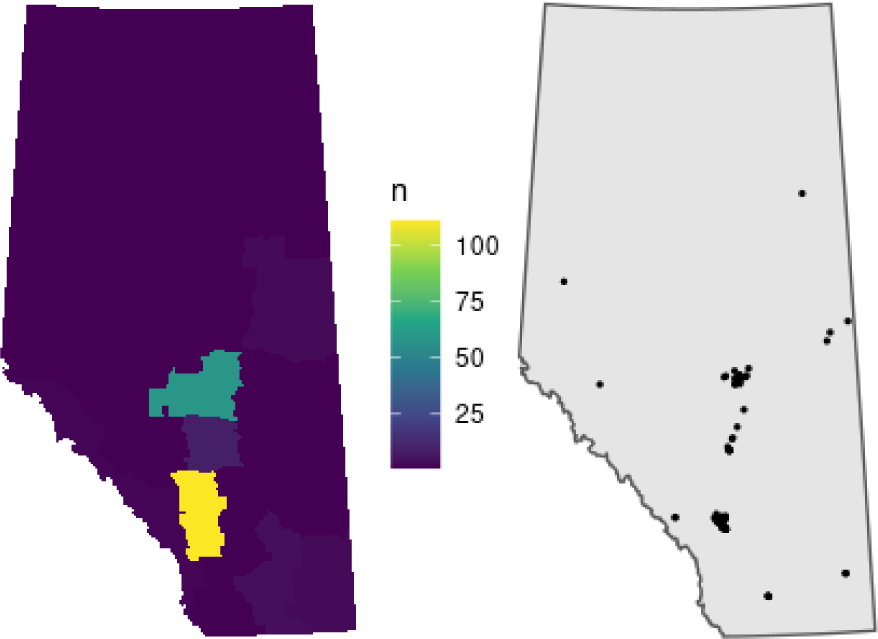
Distribution of active University of Ottawa medical school graduates in Alberta, density by census division (left) and point distribution (right).

French-speaking physicians were concentrated in Census Divisions 6 (n=464, 48.6%) and 11 (n=356, 37.3%), with only 135 (14.1%) serving the remainder of the province. Six

Census Divisions (numbers 4, 5, 7, 9, 13, and 18) had only one French-speaking physician. This distribution was similar to the general distribution of all physicians.

### Interactive Map

We also created an interactive map of Alberta-based physicians filterable by language. A draft English version is available at https://docmapper-english-test-site.netlify.app/, and bilingual maps will be published at www.docmapper.ca and www.trouvezunmedecin.ca.

## Discussion

We identified n=11,370 active physicians in the province of Alberta, of whom we further identified n=194 (1.7%) as uOttawa graduates, n=955 (8.4%) as French-speaking, and n=4,965 (43.7%) as community-based family physicians.

French-speaking physicians were concentrated in Census Division 6 (n=464, 48.6%) surrounding Calgary and Census Division 11 (n=356, 37.3%) surrounding Edmonton, with only 135 (14.1%) French-speaking physicians serving the remainder of the province. Six Census Divisions (numbers 4, 5, 7, 9, 13, and 18) had only one French-speaking physician. This distribution was similar to the general distribution of all physicians.

Overall reported French-language ability was low, with just 955 (8.4%) of all active physicians reporting competency in French. uOttawa graduates (n=70, 36.1%) were much more likely to report French ability than graduates of other schools (n=885, 7.9%), women (n=457, 9.6%) were slightly more likely than men (n=497, 7.6%), and specialists (n=666, 10.4%) were more likely than family physicians (n=289, 5.8%). These results are raw percentages reported without error bars or significance tests, because they describe an entire population or sub-population, as defined in the methods, and therefore do not constitute samples. See Appendix A for details.

### Limitations

This study has several limitations. First, it is based on a secondary analysis of data from the CPSA which is only updated annually. It is possible that our data cleaning processes categorized physicians incorrectly, either due to geocoding errors or misclassification. We were also not able to locate analyze n=3,333 physicians with no listed address and excluded them from the analysis. Finally, our uOttawa graduate analysis only includes uOttawa medical school graduates, not those who completed residency requirements or other training at uOttawa, because residency information is not available through the CPSA website. This analysis could be refined with additional data from uOttawa.

### Conclusion

In this study we have successfully collected and analyzed data about active registered physicians in the province of Alberta, Canada. We analyzed French-speaking ability and found that uOttawa graduates were much more likely the report French ability than the general population of physicians. We also produced an interactive map, filterable by language, to help patients and policymakers to find local language-concordant physicians.

This study could be extended in several directions. For example, a travel analysis could explore inequities in travel burden for French-speaking linguistic minority communities, or with additional data the analysis could expand to include uOttawa residency graduates.

## Data Availability

Physician information is available online at https://search.cpsa.ca/ . The authors' processed data is not presently available.

---

**Appendix A:** Detailed Data Tables, All Physicians.

| Characteristic (Total n=11370) | n | % |
| --- | --- | --- |
| **Gender** |  |  |
| Female | 4749 | 41.8% |
| Male | 6575 | 57.8% |
| Unknown | 46 | 0.4% |
| **Language Spoken\*** |  |  |
| English | 11370 | 100.0% |
| French | 955 | 8.4% |
| Afrikaans | 612 | 5.4% |
| Arabic | 604 | 5.3% |
| Hindi | 597 | 5.3% |
| Punjabi | 506 | 4.5% |
| Urdu | 453 | 4.0% |
| Cantonese | 266 | 2.3% |
| Spanish | 259 | 2.3% |
| Mandarin | 201 | 1.8% |
| Yoruba | 174 | 1.5% |
| German | 151 | 1.3% |
| Polish | 116 | 1.0% |
| Russian | 110 | 1.0% |
| Chinese | 105 | 0.9% |
| Gujarati | 104 | 0.9% |
| Farsi | 72 | 0.6% |
| Tamil | 65 | 0.6% |
| Romanian/Rumanian | 60 | 0.5% |
| Igbo | 59 | 0.5% |
| Italian | 59 | 0.5% |
| Bengali | 58 | 0.5% |
| Ukrainian | 57 | 0.5% |
| Portuguese | 53 | 0.5% |
| Dutch | 47 | 0.4% |
| Persian (Farsi) | 46 | 0.4% |
| Vietnamese | 38 | 0.3% |
| Korean | 36 | 0.3% |
| Hebrew | 35 | 0.3% |
| Filipino | 34 | 0.3% |
| Ibo | 33 | 0.3% |
| Malayalam | 33 | 0.3% |
| Czech | 29 | 0.3% |
| Swahili | 29 | 0.3% |
| Sindhi | 27 | 0.2% |
| Zulu | 26 | 0.2% |
| Tagalog | 24 | 0.2% |
| Telugu | 24 | 0.2% |
| Kannada | 21 | 0.2% |
| Marathi | 21 | 0.2% |
| Pashtu | 20 | 0.2% |
| Greek | 18 | 0.2% |
| Serbian | 18 | 0.2% |
| Sinhalese | 17 | 0.1% |
| Turkish | 17 | 0.1% |
| Slovak | 16 | 0.1% |
| Xhosa | 16 | 0.1% |
| Malay | 15 | 0.1% |
| Hausa | 14 | 0.1% |
| Hungarian | 14 | 0.1% |
| Croatian | 13 | 0.1% |
| Kachi | 13 | 0.1% |
| Saraiki (or Seraiki) | 13 | 0.1% |
| Japanese | 12 | 0.1% |
| Pakistani | 12 | 0.1% |
| Taiwanese | 12 | 0.1% |
| Bini | 10 | 0.1% |
| Shona | 9 | 0.1% |
| Toy-sun-ese | 9 | 0.1% |
| Kiswahili | 8 | 0.1% |
| Swedish | 8 | 0.1% |
| Kutchhi | 7 | 0.1% |
| Nepali | 7 | 0.1% |
| Serbo-Croatian | 7 | 0.1% |
| Catalan | 6 | 0.1% |
| Lithuanian | 6 | 0.1% |
| Siswati | 6 | 0.1% |
| Sotho | 6 | 0.1% |
| Amharic | 5 | 0.0% |
| Bulgarian | 5 | 0.0% |
| Norwegian | 5 | 0.0% |
| Ashanti | 4 | 0.0% |
| Burmese | 4 | 0.0% |
| Danish | 4 | 0.0% |
| Irish | 4 | 0.0% |
| Akan | 3 | 0.0% |
| Armenian | 3 | 0.0% |
| Azari | 3 | 0.0% |
| Bosnian | 3 | 0.0% |
| Gaelic | 3 | 0.0% |
| Haitian Creole | 3 | 0.0% |
| Macedonian | 3 | 0.0% |
| Mauritian | 3 | 0.0% |
| Albanian | 2 | 0.0% |
| Cebuano | 2 | 0.0% |
| Esperanto (International Language) | 2 | 0.0% |
| Finnish | 2 | 0.0% |
| Icelandic | 2 | 0.0% |
| Isoko | 2 | 0.0% |
| Setswana | 2 | 0.0% |
| Somali | 2 | 0.0% |
| Yiddish | 2 | 0.0% |
| American Sign Language | 1 | 0.0% |
| Baluchi | 1 | 0.0% |
| Basque | 1 | 0.0% |
| Bicolano | 1 | 0.0% |
| Coorgi | 1 | 0.0% |
| Cree | 1 | 0.0% |
| Fijian | 1 | 0.0% |
| Flemish | 1 | 0.0% |
| Indonesian | 1 | 0.0% |
| Kurdish (Kurmanji / Kurdi) | 1 | 0.0% |
| Maltese | 1 | 0.0% |
| Melanesian | 1 | 0.0% |
| Sidhi | 1 | 0.0% |
| Sign Language | 1 | 0.0% |
| Slovenian | 1 | 0.0% |
| Tibetan | 1 | 0.0% |
| **Physician Type** |  |  |
| Community-Based Family Physician | 4965 | 43.7% |
| Specialist | 6405 | 56.3% |
| **Practice Discipline** |  |  |
| Family Medicine | 3864 | 34.0% |
| General Practice | 1259 | 11.1% |
| Internal Medicine | 1144 | 10.1% |
| Psychiatry | 644 | 5.7% |
| Pediatrics | 594 | 5.2% |
| Diagnostic Radiology | 478 | 4.2% |
| Anesthesiology | 459 | 4.0% |
| Obstetrics | 267 | 2.3% |
| Family Medicine (Emergency Medicine) | 235 | 2.1% |
| Emergency Medicine | 233 | 2.0% |
| General Surgery | 226 | 2.0% |
| Orthopedic Surgery | 209 | 1.8% |
| Anatomical Pathology | 162 | 1.4% |
| Neurology | 147 | 1.3% |
| Ophthalmology | 139 | 1.2% |
| Physical Medicine | 109 | 1.0% |
| Dermatology | 82 | 0.7% |
| Otolaryngology - Head and Neck Surgery | 77 | 0.7% |
| Plastic Surgery | 76 | 0.7% |
| Pathology | 59 | 0.5% |
| Radiation Oncology | 57 | 0.5% |
| Urology | 57 | 0.5% |
| Cardiology | 54 | 0.5% |
| Gastroenterology | 46 | 0.4% |
| Respirology | 42 | 0.4% |
| Neurosurgery | 39 | 0.3% |
| Nephrology | 34 | 0.3% |
| Critical Care Medicine | 33 | 0.3% |
| Public Health and Preventive Medicine | 32 | 0.3% |
| Neurology (Adult and Pediatric) | 25 | 0.2% |
| Cardiac Surgery | 24 | 0.2% |
| Hematology | 22 | 0.2% |
| Medical Microbiology | 21 | 0.2% |
| Family Medicine (Palliative Care) | 20 | 0.2% |
| Community Medicine | 19 | 0.2% |
| Neonatal-perinatal Medicine | 19 | 0.2% |
| Family Medicine (Sport and Exercise Medicine) | 18 | 0.2% |
| Medical Genetics | 18 | 0.2% |
| Medical Oncology | 18 | 0.2% |
| Family Medicine (Family Practice Anesthesia) | 17 | 0.1% |
| Occupational Medicine | 15 | 0.1% |
| Family Medicine (Care of the Elderly) | 14 | 0.1% |
| Hematological Pathology | 13 | 0.1% |
| Infectious Diseases | 12 | 0.1% |
| Nuclear Medicine | 12 | 0.1% |
| Pediatric Cardiology | 12 | 0.1% |
| Pediatric Neurology | 12 | 0.1% |
| Endocrinology | 11 | 0.1% |
| Rheumatology | 10 | 0.1% |
| Family Medicine (Addiction Medicine) | 9 | 0.1% |
| Geriatric Medicine | 9 | 0.1% |
| Neurology (Pediatrics) | 9 | 0.1% |
| Vascular Surgery | 9 | 0.1% |
| Child and Adolescent Psychiatry | 7 | 0.1% |
| Neuropathology | 7 | 0.1% |
| Family Medicine (Enhanced Surgical Skills) | 6 | 0.1% |
| Pediatric Emergency Medicine | 6 | 0.1% |
| Pediatric Hematology | 6 | 0.1% |
| Clinical Immunology | 5 | 0.0% |
| Family Medicine (Obstetrical Surgical Skills) | 5 | 0.0% |
| Forensic Pathology | 5 | 0.0% |
| Forensic Psychiatry | 5 | 0.0% |
| Palliative Care | 5 | 0.0% |
| Pediatric Gastroenterology | 5 | 0.0% |
| Surgical Assists | 5 | 0.0% |
| General Internal Medicine | 4 | 0.0% |
| Geriatric Psychiatry | 4 | 0.0% |
| Maternal-Fetal Medicine | 4 | 0.0% |
| Cardiovascular | 3 | 0.0% |
| Pediatric Anesthesiology | 3 | 0.0% |
| Pediatric General Surgery | 3 | 0.0% |
| Pediatric Nephrology | 3 | 0.0% |
| Pediatrics and Pediatric Critical Care Medicine | 3 | 0.0% |
| Sport and Exercise Medicine | 3 | 0.0% |
| Transplant Surgery | 3 | 0.0% |
| Clinical Pharmacology | 2 | 0.0% |
| Hospitalist | 2 | 0.0% |
| Medical Genetics and Genomics | 2 | 0.0% |
| Palliative Medicine | 2 | 0.0% |
| Pediatric Endocrinology | 2 | 0.0% |
| Pediatric Intensive Care | 2 | 0.0% |
| Pediatric Neonatal-perinatal Medicine | 2 | 0.0% |
| Pediatric Respiriology | 2 | 0.0% |
| Pediatric Rheumatology | 2 | 0.0% |
| Pediatric Urology | 2 | 0.0% |
| Pediatrics and Pediatric Emergency Medicine | 2 | 0.0% |
| Abdominal Wall Hernia Consultation | 1 | 0.0% |
| Adolescent Medicine | 1 | 0.0% |
| Anesthesia and Pediatric Critical Care Medicine | 1 | 0.0% |
| British Army Training Unit | 1 | 0.0% |
| CAC (Enhanced Surgical Skills) | 1 | 0.0% |
| CAC (Family Practice Anesthesia) | 1 | 0.0% |
| CAC (Obstetrical Surgical Skills) | 1 | 0.0% |
| Clinical Assistant | 1 | 0.0% |
| Developmental Pediatrics | 1 | 0.0% |
| General and Anatomical Pathology | 1 | 0.0% |
| General Surgical Oncology | 1 | 0.0% |
| Medical management of neurosurgical patients | 1 | 0.0% |
| Medical Ophthalmology | 1 | 0.0% |
| Neonatal-Perinatal Medicine | 1 | 0.0% |
| Neuroradiology | 1 | 0.0% |
| Pain Management | 1 | 0.0% |
| Pediatric Critical Care Medicine | 1 | 0.0% |
| Pediatric Orthopedic Surgery and the care of adults as part of the Complex Spinal Deformity Program | 1 | 0.0% |
| Pediatric Radiology | 1 | 0.0% |
| Pediatric Surgery | 1 | 0.0% |
| Pediatrics and Infectious Disease | 1 | 0.0% |
| Rheumatology and Clinical Immunology | 1 | 0.0% |
| Telemedicine | 1 | 0.0% |
| **Institution / Country of Degree** |  |  |
| UofA - University of Alberta | 2787 | 24.5% |
| UofC - University of Calgary | 1694 | 14.9% |
| South Africa | 747 | 6.6% |
| University of British Columbia | 485 | 4.3% |
| University of Saskatchewan | 442 | 3.9% |
| Nigeria | 392 | 3.4% |
| University of Manitoba | 324 | 2.8% |
| University of Toronto | 324 | 2.8% |
| India | 292 | 2.6% |
| Pakistan | 281 | 2.5% |
| Libya | 226 | 2.0% |
| Queen's University | 223 | 2.0% |
| University of Western Ontario | 199 | 1.8% |
| University of Ottawa | 194 | 1.7% |
| McMaster University | 193 | 1.7% |
| Dalhousie University | 191 | 1.7% |
| McGill University | 163 | 1.4% |
| United Kingdom | 163 | 1.4% |
| Ireland | 145 | 1.3% |
| Egypt | 140 | 1.2% |
| Memorial University | 131 | 1.2% |
| United States of America | 92 | 0.8% |
| Australia | 83 | 0.7% |
| Iran | 83 | 0.7% |
| England | 77 | 0.7% |
| Iraq | 77 | 0.7% |
| Poland | 50 | 0.4% |
| Saudi Arabia | 49 | 0.4% |
| Philippines | 42 | 0.4% |
| Romania | 42 | 0.4% |
| China | 38 | 0.3% |
| Universite de Montreal | 34 | 0.3% |
| Grenada | 33 | 0.3% |
| Germany | 32 | 0.3% |
| Scotland | 32 | 0.3% |
| Colombia | 31 | 0.3% |
| Russia | 30 | 0.3% |
| Universite de Sherbrooke | 28 | 0.2% |
| Sudan | 26 | 0.2% |
| Universite Laval | 26 | 0.2% |
| Brazil | 25 | 0.2% |
| Netherlands Antilles | 25 | 0.2% |
| Bangladesh | 23 | 0.2% |
| Saint Kitts and Nevis | 23 | 0.2% |
| Syria | 23 | 0.2% |
| Ukraine | 23 | 0.2% |
| The Netherlands | 19 | 0.2% |
| Barbados | 18 | 0.2% |
| Mexico | 17 | 0.1% |
| China China | 16 | 0.1% |
| Northern Ontario School of Medicine | 16 | 0.1% |
| Poland Poland | 16 | 0.1% |
| Argentina | 15 | 0.1% |
| Hungary | 15 | 0.1% |
| Sri Lanka | 14 | 0.1% |
| Uganda | 14 | 0.1% |
| Italy | 13 | 0.1% |
| Saba | 13 | 0.1% |
| Zimbabwe | 13 | 0.1% |
| Spain | 12 | 0.1% |
| Turkey | 12 | 0.1% |
| Democratic Republic of the Congo | 10 | 0.1% |
| Dominica | 10 | 0.1% |
| Israel | 10 | 0.1% |
| Cayman Islands | 9 | 0.1% |
| Ghana | 9 | 0.1% |
| Hong Kong | 9 | 0.1% |
| Mexico Mexico | 9 | 0.1% |
| Moldova, Republic of | 9 | 0.1% |
| New Zealand | 9 | 0.1% |
| Slovakia | 9 | 0.1% |
| Antigua and Barbuda | 8 | 0.1% |
| Czechoslovakia | 8 | 0.1% |
| Trinidad | 8 | 0.1% |
| Lebanon | 7 | 0.1% |
| Northern Ireland | 7 | 0.1% |
| Serbia | 7 | 0.1% |
| Venezuela | 7 | 0.1% |
| Yugoslavia | 7 | 0.1% |
| France | 6 | 0.1% |
| Jordan | 6 | 0.1% |
| Russia Russia | 6 | 0.1% |
| Austria | 5 | 0.0% |
| Belgium | 5 | 0.0% |
| Dominican Republic | 5 | 0.0% |
| United Arab Emirates | 5 | 0.0% |
| West Indies | 5 | 0.0% |
| Bahrain | 4 | 0.0% |
| Belarus | 4 | 0.0% |
| Cuba | 4 | 0.0% |
| Czecho-Slovakia Czech Republic | 4 | 0.0% |
| German Germany | 4 | 0.0% |
| Jamaica | 4 | 0.0% |
| Kenya | 4 | 0.0% |
| Malaysia | 4 | 0.0% |
| Singapore | 4 | 0.0% |
| Zambia | 4 | 0.0% |
| Algeria | 3 | 0.0% |
| Chile | 3 | 0.0% |
| Czech Republic | 3 | 0.0% |
| Greece | 3 | 0.0% |
| Korea | 3 | 0.0% |
| Lithuania | 3 | 0.0% |
| Morocco | 3 | 0.0% |
| Peru | 3 | 0.0% |
| Physician Lithuania | 3 | 0.0% |
| Saint Lucia | 3 | 0.0% |
| Saint Vincent and the Grenadines | 3 | 0.0% |
| Sint Maarten | 3 | 0.0% |
| Tanzania | 3 | 0.0% |
| Wales | 3 | 0.0% |
| Yemen | 3 | 0.0% |
| Albania | 2 | 0.0% |
| Aruba | 2 | 0.0% |
| Belize | 2 | 0.0% |
| Bosnia | 2 | 0.0% |
| Bosnia Bosnia - Herzegovina | 2 | 0.0% |
| Costa Rica | 2 | 0.0% |
| El Salvador | 2 | 0.0% |
| Ethiopia | 2 | 0.0% |
| Holland | 2 | 0.0% |
| Japan | 2 | 0.0% |
| Kazakhstan | 2 | 0.0% |
| Mexico Bolivia | 2 | 0.0% |
| Nepal | 2 | 0.0% |
| Nicaragua | 2 | 0.0% |
| Physician Bulgaria | 2 | 0.0% |
| Spain Spain | 2 | 0.0% |
| Switzerland | 2 | 0.0% |
| Viet Nam | 2 | 0.0% |
| Afghanistan | 1 | 0.0% |
| Anguilla | 1 | 0.0% |
| Armenia | 1 | 0.0% |
| Azerbaijan | 1 | 0.0% |
| Azerbaijan Azerbaijan | 1 | 0.0% |
| Bolivia | 1 | 0.0% |
| Bosnia Bosnia | 1 | 0.0% |
| Bulgaria | 1 | 0.0% |
| Burma | 1 | 0.0% |
| China Australia | 1 | 0.0% |
| China England | 1 | 0.0% |
| China Taiwan | 1 | 0.0% |
| Croatia | 1 | 0.0% |
| Curacao | 1 | 0.0% |
| Czecho-Slovakia Slovakia | 1 | 0.0% |
| Denmark | 1 | 0.0% |
| Estonia | 1 | 0.0% |
| Fiji | 1 | 0.0% |
| Finland | 1 | 0.0% |
| Guatemala | 1 | 0.0% |
| Guyana | 1 | 0.0% |
| Iceland | 1 | 0.0% |
| Kuwait | 1 | 0.0% |
| Kyrgyzstan | 1 | 0.0% |
| Liberia | 1 | 0.0% |
| Mexico Colombia | 1 | 0.0% |
| Mexico Peru | 1 | 0.0% |
| Mongolia | 1 | 0.0% |
| Myanmar | 1 | 0.0% |
| ONTARIO Conversion | 1 | 0.0% |
| Physician Albania | 1 | 0.0% |
| Physician Holland | 1 | 0.0% |
| Physician The Netherlands | 1 | 0.0% |
| Portugal Portugal | 1 | 0.0% |
| Republic of Congo | 1 | 0.0% |
| Rhodesia | 1 | 0.0% |
| Russia Kyrgyzstan | 1 | 0.0% |
| Somalia | 1 | 0.0% |
| Sultanate of Oman | 1 | 0.0% |
| Sweden | 1 | 0.0% |
| Sweden Sweden | 1 | 0.0% |
| Taiwan | 1 | 0.0% |
| Thailand | 1 | 0.0% |
| UofC - Undergraduate Elective | 1 | 0.0% |
| UofC - University of Calgary Undergrads | 1 | 0.0% |
| Uruguay | 1 | 0.0% |
| West Germany | 1 | 0.0% |
| Zaire | 1 | 0.0% |

\* Respondents could report competency in more than one language.

**Appendix B:**
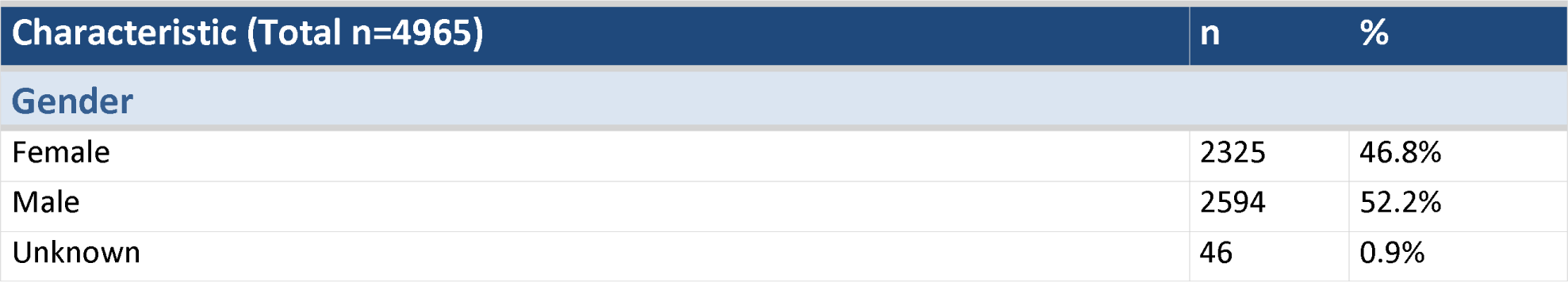
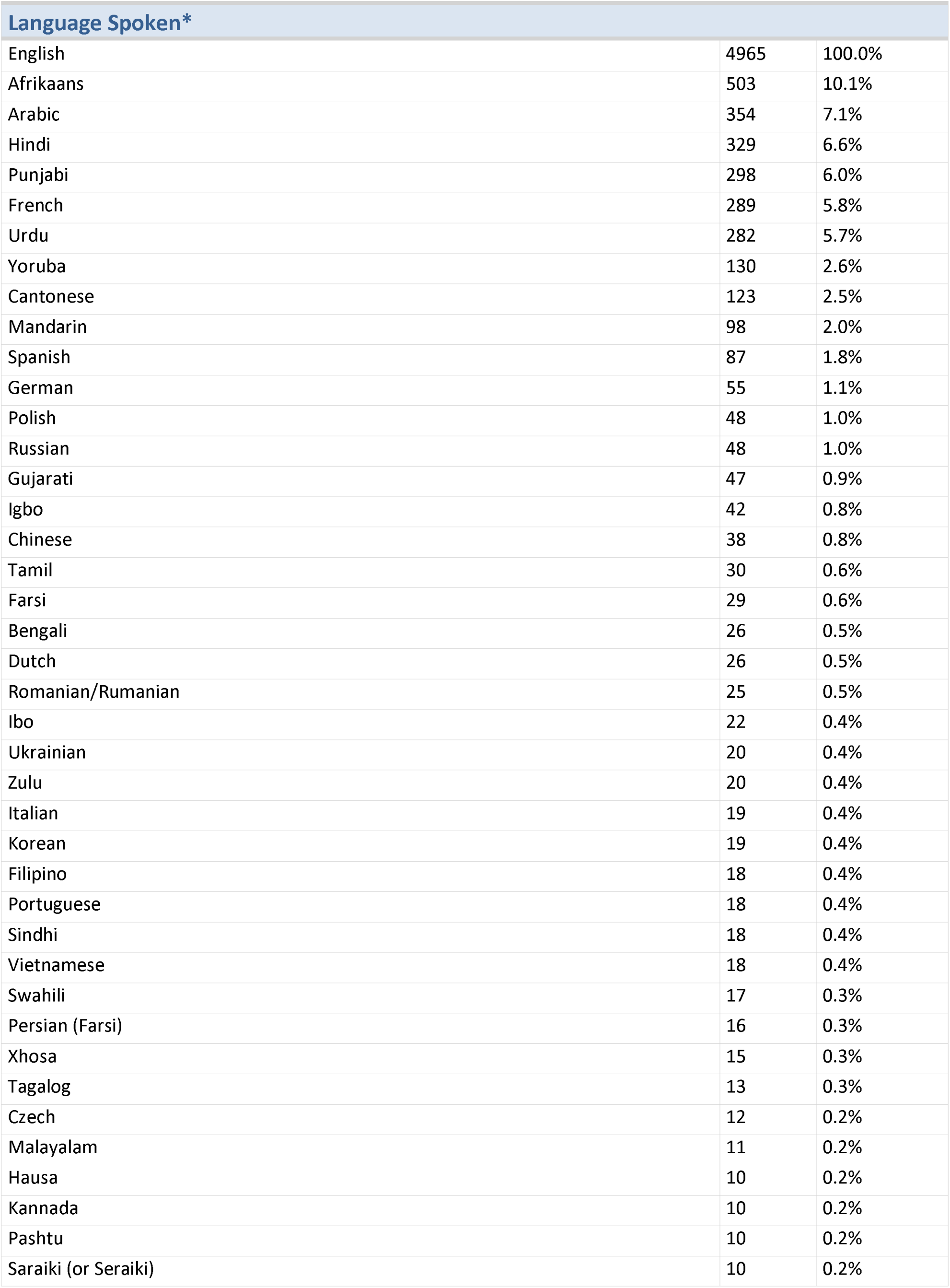
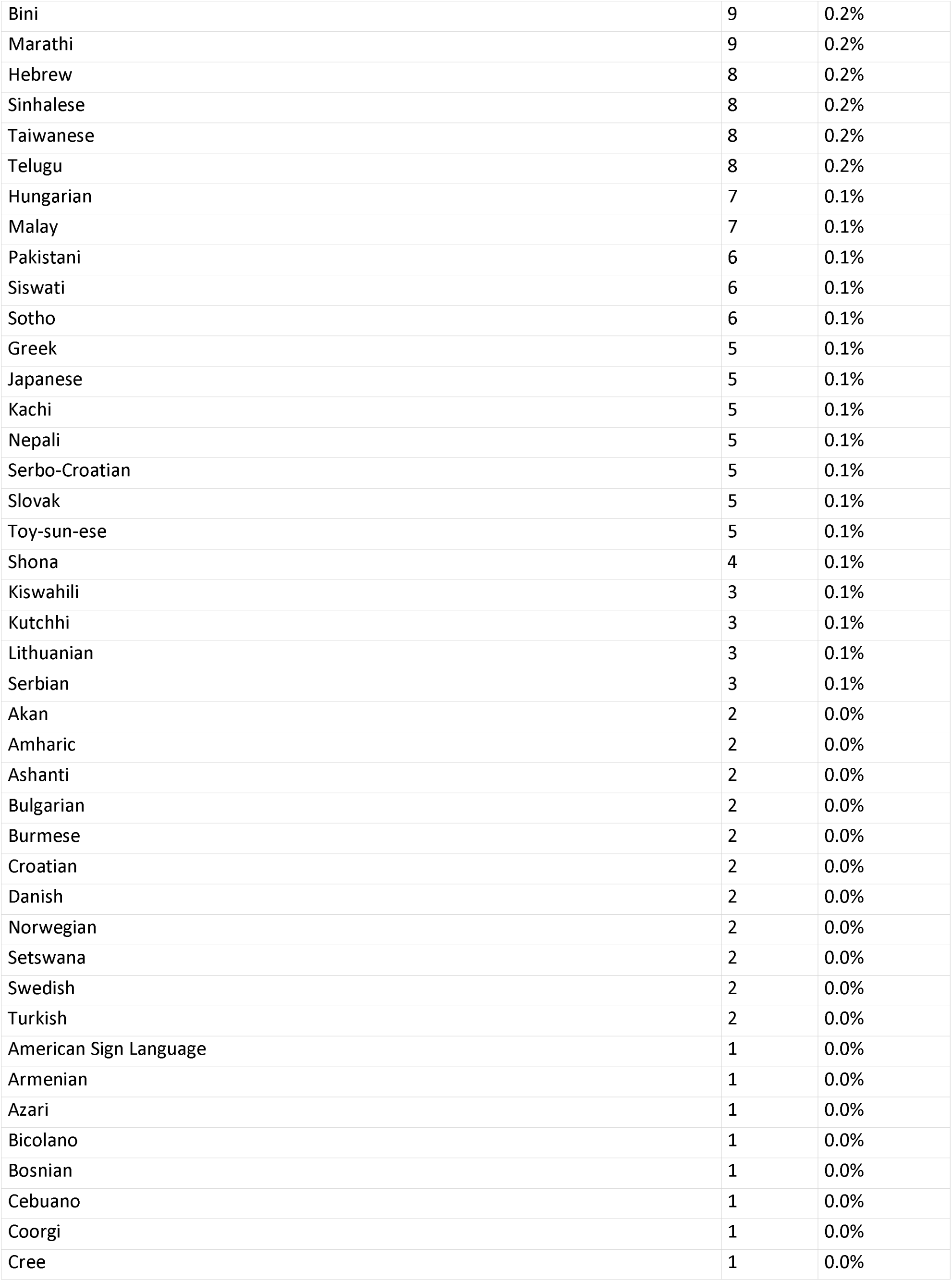
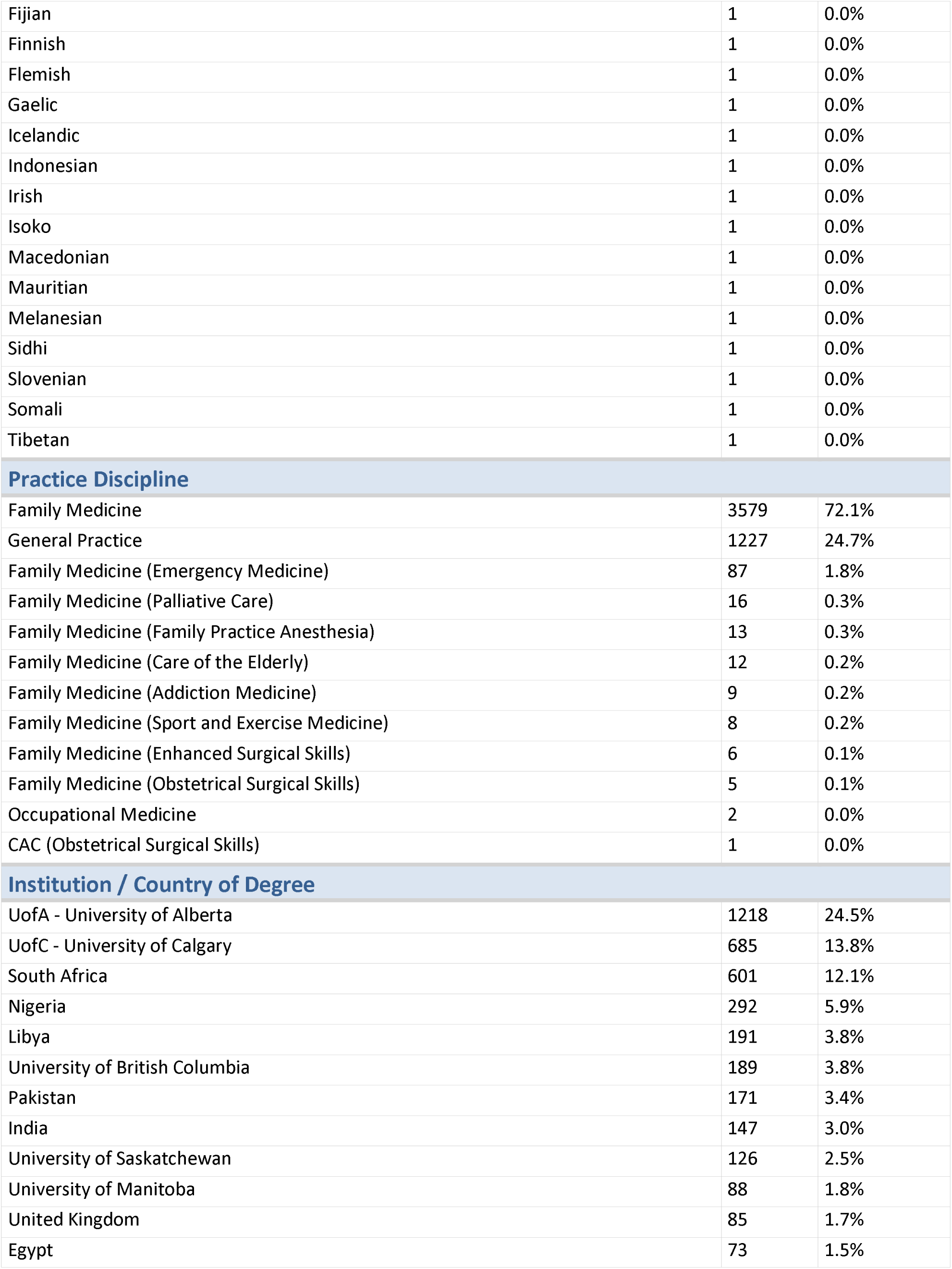
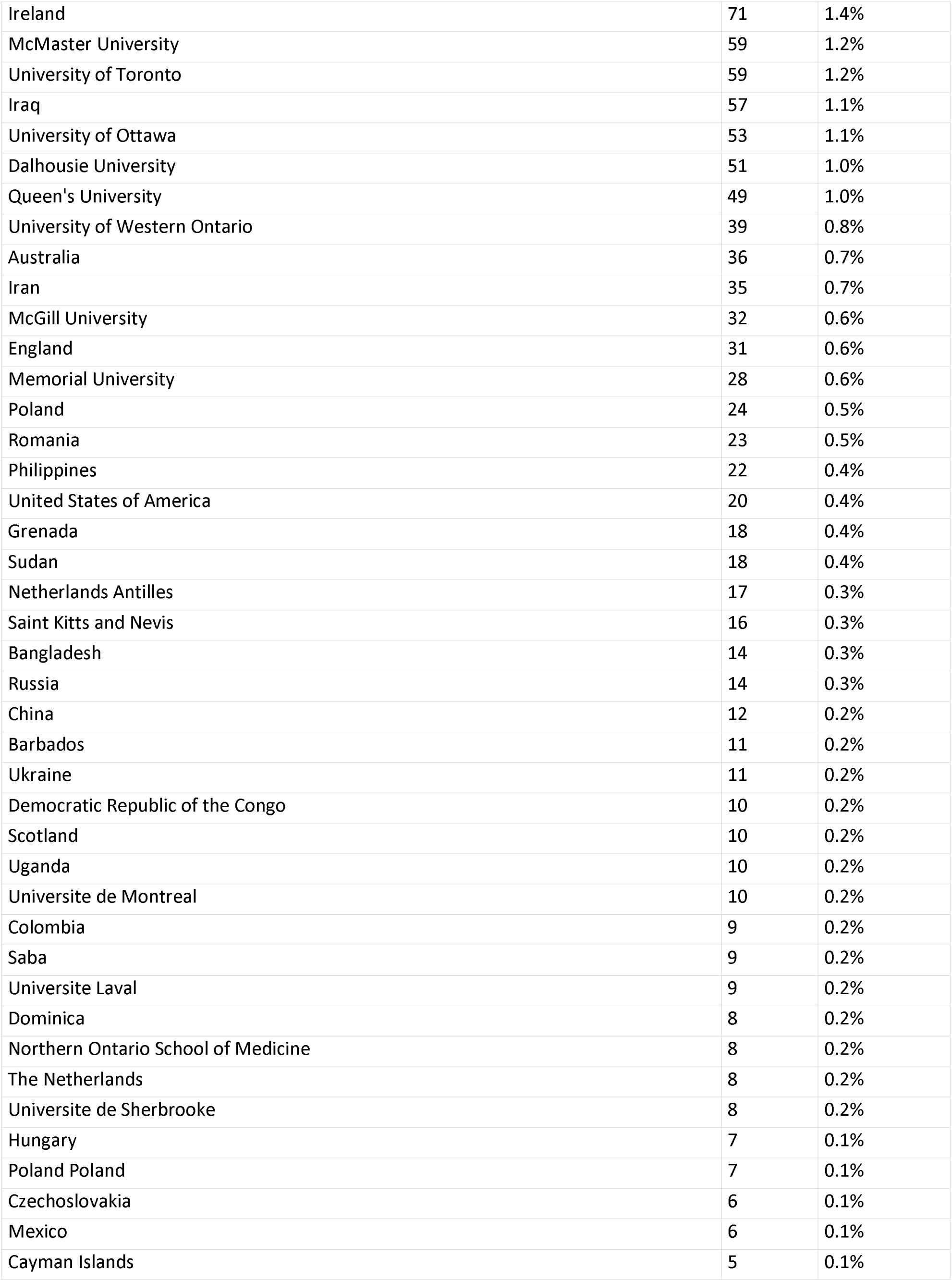
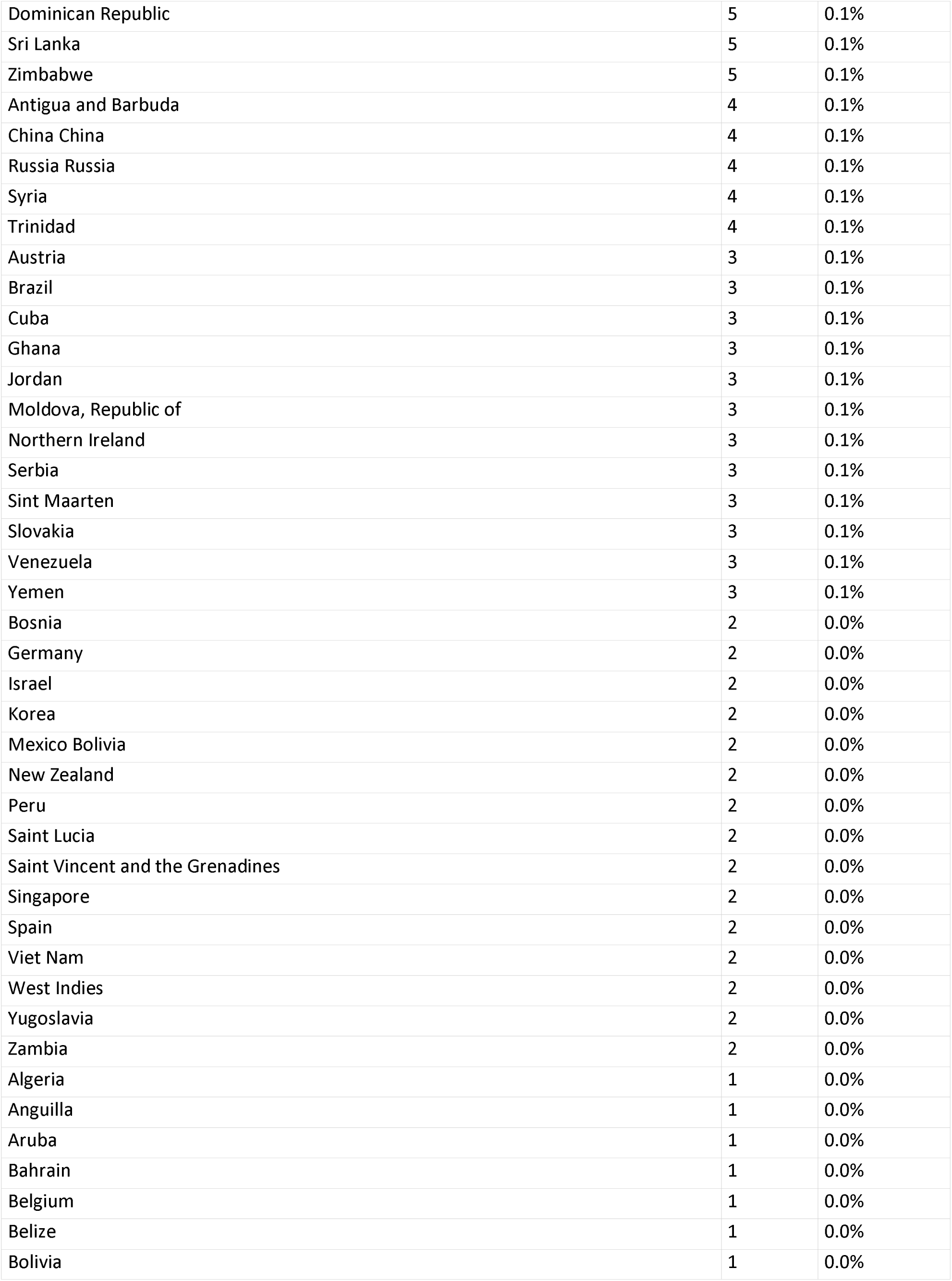
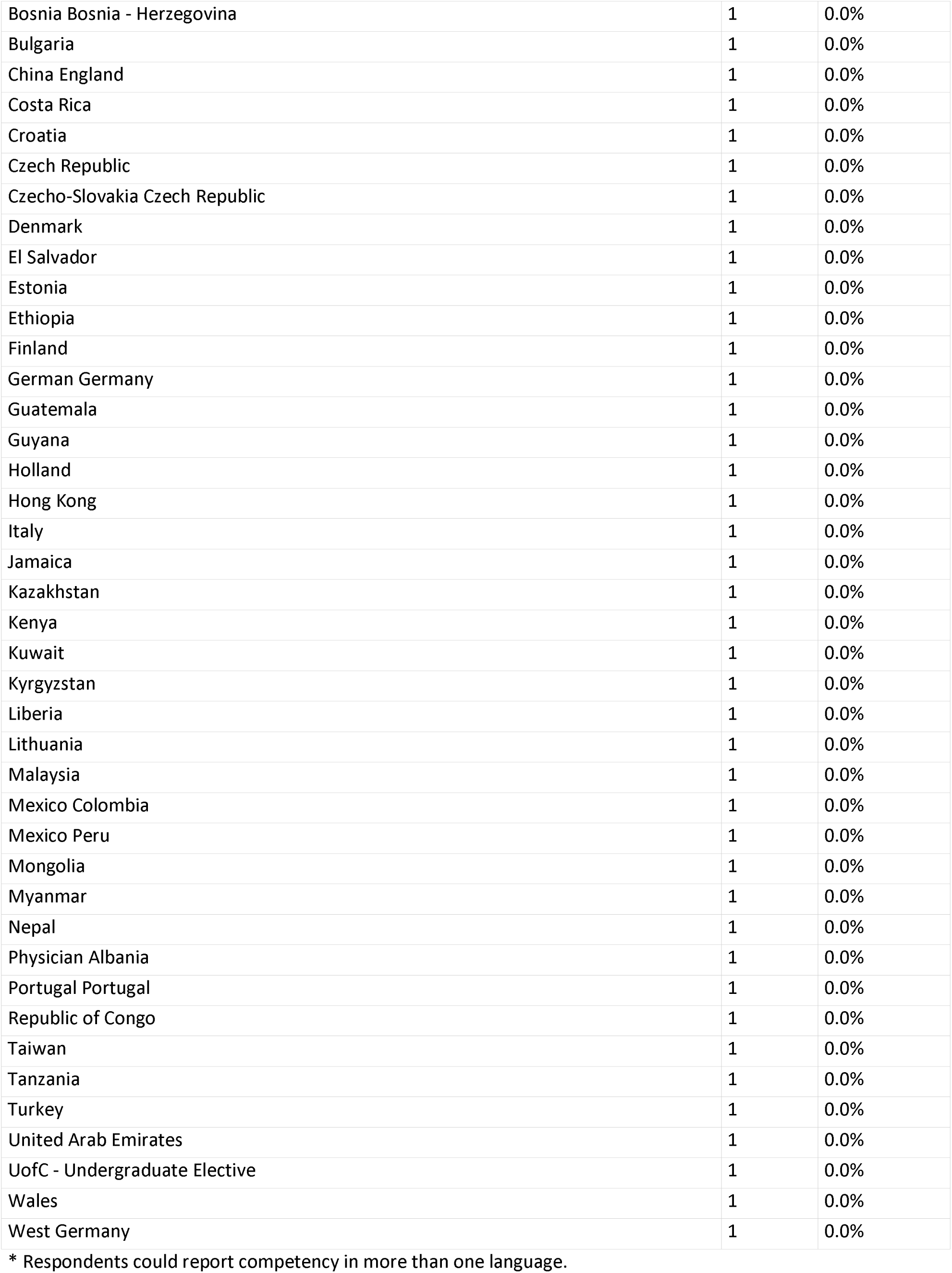
Detailed Data Tables, Family Physicians.

| Characteristic (Total n=4965) | n | % |
| --- | --- | --- |
| **Gender** |  |  |
| Female | 2325 | 46.8% |
| Male | 2594 | 52.2% |
| Unknown | 46 | 0.9% |

| Language Spoken* |  |  |
| --- | --- | --- |
| English | 4965 | 100.0% |
| Afrikaans | 503 | 10.1% |
| Arabic | 354 | 7.1% |
| Hindi | 329 | 6.6% |
| Punjabi | 298 | 6.0% |
| French | 289 | 5.8% |
| Urdu | 282 | 5.7% |
| Yoruba | 130 | 2.6% |
| Cantonese | 123 | 2.5% |
| Mandarin | 98 | 2.0% |
| Spanish | 87 | 1.8% |
| German | 55 | 1.1% |
| Polish | 48 | 1.0% |
| Russian | 48 | 1.0% |
| Gujarati | 47 | 0.9% |
| Igbo | 42 | 0.8% |
| Chinese | 38 | 0.8% |
| Tamil | 30 | 0.6% |
| Farsi | 29 | 0.6% |
| Bengali | 26 | 0.5% |
| Dutch | 26 | 0.5% |
| Romanian/Rumanian | 25 | 0.5% |
| Ibo | 22 | 0.4% |
| Ukrainian | 20 | 0.4% |
| Zulu | 20 | 0.4% |
| Italian | 19 | 0.4% |
| Korean | 19 | 0.4% |
| Filipino | 18 | 0.4% |
| Portuguese | 18 | 0.4% |
| Sindhi | 18 | 0.4% |
| Vietnamese | 18 | 0.4% |
| Swahili | 17 | 0.3% |
| Persian (Farsi) | 16 | 0.3% |
| Xhosa | 15 | 0.3% |
| Tagalog | 13 | 0.3% |
| Czech | 12 | 0.2% |
| Malayalam | 11 | 0.2% |
| Hausa | 10 | 0.2% |
| Kannada | 10 | 0.2% |
| Pashtu | 10 | 0.2% |
| Saraiki (or Seraiki) | 10 | 0.2% |
| Bini | 9 | 0.2% |
| Marathi | 9 | 0.2% |
| Hebrew | 8 | 0.2% |
| Sinhalese | 8 | 0.2% |
| Taiwanese | 8 | 0.2% |
| Telugu | 8 | 0.2% |
| Hungarian | 7 | 0.1% |
| Malay | 7 | 0.1% |
| Pakistani | 6 | 0.1% |
| Siswati | 6 | 0.1% |
| Sotho | 6 | 0.1% |
| Greek | 5 | 0.1% |
| Japanese | 5 | 0.1% |
| Kachi | 5 | 0.1% |
| Nepali | 5 | 0.1% |
| Serbo-Croatian | 5 | 0.1% |
| Slovak | 5 | 0.1% |
| Toy-sun-ese | 5 | 0.1% |
| Shona | 4 | 0.1% |
| Kiswahili | 3 | 0.1% |
| Kutchhi | 3 | 0.1% |
| Lithuanian | 3 | 0.1% |
| Serbian | 3 | 0.1% |
| Akan | 2 | 0.0% |
| Amharic | 2 | 0.0% |
| Ashanti | 2 | 0.0% |
| Bulgarian | 2 | 0.0% |
| Burmese | 2 | 0.0% |
| Croatian | 2 | 0.0% |
| Danish | 2 | 0.0% |
| Norwegian | 2 | 0.0% |
| Setswana | 2 | 0.0% |
| Swedish | 2 | 0.0% |
| Turkish | 2 | 0.0% |
| American Sign Language | 1 | 0.0% |
| Armenian | 1 | 0.0% |
| Azari | 1 | 0.0% |
| Bicolano | 1 | 0.0% |
| Bosnian | 1 | 0.0% |
| Cebuano | 1 | 0.0% |
| Coorgi | 1 | 0.0% |
| Cree | 1 | 0.0% |
| Fijian | 1 | 0.0% |
| Finnish | 1 | 0.0% |
| Flemish | 1 | 0.0% |
| Gaelic | 1 | 0.0% |
| Icelandic | 1 | 0.0% |
| Indonesian | 1 | 0.0% |
| Irish | 1 | 0.0% |
| Isoko | 1 | 0.0% |
| Macedonian | 1 | 0.0% |
| Mauritian | 1 | 0.0% |
| Melanesian | 1 | 0.0% |
| Sidhi | 1 | 0.0% |
| Slovenian | 1 | 0.0% |
| Somali | 1 | 0.0% |
| Tibetan | 1 | 0.0% |
| **Practice Discipline** |  |  |
| Family Medicine | 3579 | 72.1% |
| General Practice | 1227 | 24.7% |
| Family Medicine (Emergency Medicine) | 87 | 1.8% |
| Family Medicine (Palliative Care) | 16 | 0.3% |
| Family Medicine (Family Practice Anesthesia) | 13 | 0.3% |
| Family Medicine (Care of the Elderly) | 12 | 0.2% |
| Family Medicine (Addiction Medicine) | 9 | 0.2% |
| Family Medicine (Sport and Exercise Medicine) | 8 | 0.2% |
| Family Medicine (Enhanced Surgical Skills) | 6 | 0.1% |
| Family Medicine (Obstetrical Surgical Skills) | 5 | 0.1% |
| Occupational Medicine | 2 | 0.0% |
| CAC (Obstetrical Surgical Skills) | 1 | 0.0% |
| **Institution / Country of Degree** |  |  |
| UofA - University of Alberta | 1218 | 24.5% |
| UofC - University of Calgary | 685 | 13.8% |
| South Africa | 601 | 12.1% |
| Nigeria | 292 | 5.9% |
| Libya | 191 | 3.8% |
| University of British Columbia | 189 | 3.8% |
| Pakistan | 171 | 3.4% |
| India | 147 | 3.0% |
| University of Saskatchewan | 126 | 2.5% |
| University of Manitoba | 88 | 1.8% |
| United Kingdom | 85 | 1.7% |
| Egypt | 73 | 1.5% |
| Ireland | 71 | 1.4% |
| McMaster University | 59 | 1.2% |
| University of Toronto | 59 | 1.2% |
| Iraq | 57 | 1.1% |
| University of Ottawa | 53 | 1.1% |
| Dalhousie University | 51 | 1.0% |
| Queen's University | 49 | 1.0% |
| University of Western Ontario | 39 | 0.8% |
| Australia | 36 | 0.7% |
| Iran | 35 | 0.7% |
| McGill University | 32 | 0.6% |
| England | 31 | 0.6% |
| Memorial University | 28 | 0.6% |
| Poland | 24 | 0.5% |
| Romania | 23 | 0.5% |
| Philippines | 22 | 0.4% |
| United States of America | 20 | 0.4% |
| Grenada | 18 | 0.4% |
| Sudan | 18 | 0.4% |
| Netherlands Antilles | 17 | 0.3% |
| Saint Kitts and Nevis | 16 | 0.3% |
| Bangladesh | 14 | 0.3% |
| Russia | 14 | 0.3% |
| China | 12 | 0.2% |
| Barbados | 11 | 0.2% |
| Ukraine | 11 | 0.2% |
| Democratic Republic of the Congo | 10 | 0.2% |
| Scotland | 10 | 0.2% |
| Uganda | 10 | 0.2% |
| Universite de Montreal | 10 | 0.2% |
| Colombia | 9 | 0.2% |
| Saba | 9 | 0.2% |
| Universite Laval | 9 | 0.2% |
| Dominica | 8 | 0.2% |
| Northern Ontario School of Medicine | 8 | 0.2% |
| The Netherlands | 8 | 0.2% |
| Universite de Sherbrooke | 8 | 0.2% |
| Hungary | 7 | 0.1% |
| Poland Poland | 7 | 0.1% |
| Czechoslovakia | 6 | 0.1% |
| Mexico | 6 | 0.1% |
| Cayman Islands | 5 | 0.1% |
| Dominican Republic | 5 | 0.1% |
| Sri Lanka | 5 | 0.1% |
| Zimbabwe | 5 | 0.1% |
| Antigua and Barbuda | 4 | 0.1% |
| China China | 4 | 0.1% |
| Russia Russia | 4 | 0.1% |
| Syria | 4 | 0.1% |
| Trinidad | 4 | 0.1% |
| Austria | 3 | 0.1% |
| Brazil | 3 | 0.1% |
| Cuba | 3 | 0.1% |
| Ghana | 3 | 0.1% |
| Jordan | 3 | 0.1% |
| Moldova, Republic of | 3 | 0.1% |
| Northern Ireland | 3 | 0.1% |
| Serbia | 3 | 0.1% |
| Sint Maarten | 3 | 0.1% |
| Slovakia | 3 | 0.1% |
| Venezuela | 3 | 0.1% |
| Yemen | 3 | 0.1% |
| Bosnia | 2 | 0.0% |
| Germany | 2 | 0.0% |
| Israel | 2 | 0.0% |
| Korea | 2 | 0.0% |
| Mexico Bolivia | 2 | 0.0% |
| New Zealand | 2 | 0.0% |
| Peru | 2 | 0.0% |
| Saint Lucia | 2 | 0.0% |
| Saint Vincent and the Grenadines | 2 | 0.0% |
| Singapore | 2 | 0.0% |
| Spain | 2 | 0.0% |
| Viet Nam | 2 | 0.0% |
| West Indies | 2 | 0.0% |
| Yugoslavia | 2 | 0.0% |
| Zambia | 2 | 0.0% |
| Algeria | 1 | 0.0% |
| Anguilla | 1 | 0.0% |
| Aruba | 1 | 0.0% |
| Bahrain | 1 | 0.0% |
| Belgium | 1 | 0.0% |
| Belize | 1 | 0.0% |
| Bolivia | 1 | 0.0% |
| Bosnia Bosnia - Herzegovina | 1 | 0.0% |
| Bulgaria | 1 | 0.0% |
| China England | 1 | 0.0% |
| Costa Rica | 1 | 0.0% |
| Croatia | 1 | 0.0% |
| Czech Republic | 1 | 0.0% |
| Czecho-Slovakia Czech Republic | 1 | 0.0% |
| Denmark | 1 | 0.0% |
| El Salvador | 1 | 0.0% |
| Estonia | 1 | 0.0% |
| Ethiopia | 1 | 0.0% |
| Finland | 1 | 0.0% |
| German Germany | 1 | 0.0% |
| Guatemala | 1 | 0.0% |
| Guyana | 1 | 0.0% |
| Holland | 1 | 0.0% |
| Hong Kong | 1 | 0.0% |
| Italy | 1 | 0.0% |
| Jamaica | 1 | 0.0% |
| Kazakhstan | 1 | 0.0% |
| Kenya | 1 | 0.0% |
| Kuwait | 1 | 0.0% |
| Kyrgyzstan | 1 | 0.0% |
| Liberia | 1 | 0.0% |
| Lithuania | 1 | 0.0% |
| Malaysia | 1 | 0.0% |
| Mexico Colombia | 1 | 0.0% |
| Mexico Peru | 1 | 0.0% |
| Mongolia | 1 | 0.0% |
| Myanmar | 1 | 0.0% |
| Nepal | 1 | 0.0% |
| Physician Albania | 1 | 0.0% |
| Portugal Portugal | 1 | 0.0% |
| Republic of Congo | 1 | 0.0% |
| Taiwan | 1 | 0.0% |
| Tanzania | 1 | 0.0% |
| Turkey | 1 | 0.0% |
| United Arab Emirates | 1 | 0.0% |
| UofC - Undergraduate Elective | 1 | 0.0% |
| Wales | 1 | 0.0% |
| West Germany | 1 | 0.0% |

\* Respondents could report competency in more than one language.

**Appendix C:** Detailed Data Tables, French-Speaking Physicians.

| Characteristic (Total n=955) | n | % |
| --- | --- | --- |
| **Gender** |  |  |
| Female | 457 | 47.9% |
| Male | 497 | 52.0% |
| Unknown | 1 | 0.1% |
| **Language Spoken\*** |  |  |
| English | 955 | 100.0% |
| French | 955 | 100.0% |
| Spanish | 68 | 7.1% |
| Arabic | 43 | 4.5% |
| German | 41 | 4.3% |
| Hindi | 19 | 2.0% |
| Italian | 16 | 1.7% |
| Polish | 16 | 1.7% |
| Romanian/Rumanian | 15 | 1.6% |
| Cantonese | 13 | 1.4% |
| Mandarin | 11 | 1.2% |
| Gujarati | 8 | 0.8% |
| Farsi | 7 | 0.7% |
| Swahili | 7 | 0.7% |
| Urdu | 7 | 0.7% |
| Czech | 6 | 0.6% |
| Dutch | 6 | 0.6% |
| Hebrew | 6 | 0.6% |
| Bengali | 5 | 0.5% |
| Chinese | 5 | 0.5% |
| Greek | 5 | 0.5% |
| Portuguese | 5 | 0.5% |
| Russian | 5 | 0.5% |
| Ukrainian | 5 | 0.5% |
| Tamil | 4 | 0.4% |
| Afrikaans | 3 | 0.3% |
| Haitian Creole | 3 | 0.3% |
| Kachi | 3 | 0.3% |
| Mauritian | 3 | 0.3% |
| Punjabi | 3 | 0.3% |
| Serbian | 3 | 0.3% |
| Vietnamese | 3 | 0.3% |
| Japanese | 2 | 0.2% |
| Korean | 2 | 0.2% |
| Lithuanian | 2 | 0.2% |
| Yiddish | 2 | 0.2% |
| American Sign Language | 1 | 0.1% |
| Amharic | 1 | 0.1% |
| Ashanti | 1 | 0.1% |
| Catalan | 1 | 0.1% |
| Croatian | 1 | 0.1% |
| Esperanto (International Language) | 1 | 0.1% |
| Filipino | 1 | 0.1% |
| Gaelic | 1 | 0.1% |
| Hungarian | 1 | 0.1% |
| Icelandic | 1 | 0.1% |
| Kiswahili | 1 | 0.1% |
| Macedonian | 1 | 0.1% |
| Marathi | 1 | 0.1% |
| Norwegian | 1 | 0.1% |
| Serbo-Croatian | 1 | 0.1% |
| Somali | 1 | 0.1% |
| Sotho | 1 | 0.1% |
| Swedish | 1 | 0.1% |
| Tagalog | 1 | 0.1% |
| Taiwanese | 1 | 0.1% |
| Zulu | 1 | 0.1% |
| **Physician Type** |  |  |
| Community-Based Family Physician | 289 | 30.3% |
| Specialist | 666 | 69.7% |
| **Practice Discipline** |  |  |
| Family Medicine | 264 | 27.6% |
| Internal Medicine | 113 | 11.8% |
| Pediatrics | 71 | 7.4% |
| Anesthesiology | 51 | 5.3% |
| General Practice | 49 | 5.1% |
| Diagnostic Radiology | 46 | 4.8% |
| Orthopedic Surgery | 35 | 3.7% |
| Obstetrics | 34 | 3.6% |
| Psychiatry | 32 | 3.4% |
| General Surgery | 28 | 2.9% |
| Emergency Medicine | 26 | 2.7% |
| Family Medicine (Emergency Medicine) | 22 | 2.3% |
| Neurology | 18 | 1.9% |
| Ophthalmology | 16 | 1.7% |
| Anatomical Pathology | 15 | 1.6% |
| Dermatology | 9 | 0.9% |
| Otolaryngology - Head and Neck Surgery | 8 | 0.8% |
| Plastic Surgery | 8 | 0.8% |
| Nephrology | 7 | 0.7% |
| Critical Care Medicine | 6 | 0.6% |
| Medical Genetics | 6 | 0.6% |
| Physical Medicine | 6 | 0.6% |
| Radiation Oncology | 6 | 0.6% |
| Neurosurgery | 5 | 0.5% |
| Respirology | 5 | 0.5% |
| Urology | 5 | 0.5% |
| Cardiology | 4 | 0.4% |
| Gastroenterology | 4 | 0.4% |
| Infectious Diseases | 4 | 0.4% |
| Public Health and Preventive Medicine | 4 | 0.4% |
| Family Medicine (Sport and Exercise Medicine) | 3 | 0.3% |
| Hematology | 3 | 0.3% |
| Medical Oncology | 3 | 0.3% |
| Pathology | 3 | 0.3% |
| Pediatric Neurology | 3 | 0.3% |
| Endocrinology | 2 | 0.2% |
| Family Medicine (Addiction Medicine) | 2 | 0.2% |
| Neurology (Pediatrics) | 2 | 0.2% |
| Nuclear Medicine | 2 | 0.2% |
| Pediatric Cardiology | 2 | 0.2% |
| Pediatric Hematology | 2 | 0.2% |
| Vascular Surgery | 2 | 0.2% |
| Adolescent Medicine | 1 | 0.1% |
| CAC (Enhanced Surgical Skills) | 1 | 0.1% |
| Cardiac Surgery | 1 | 0.1% |
| Clinical Immunology | 1 | 0.1% |
| Family Medicine (Care of the Elderly) | 1 | 0.1% |
| Family Medicine (Obstetrical Surgical Skills) | 1 | 0.1% |
| Forensic Psychiatry | 1 | 0.1% |
| Medical Microbiology | 1 | 0.1% |
| Neonatal-perinatal Medicine | 1 | 0.1% |
| Neurology (Adult and Pediatric) | 1 | 0.1% |
| Occupational Medicine | 1 | 0.1% |
| Palliative Care | 1 | 0.1% |
| Pediatric Emergency Medicine | 1 | 0.1% |
| Pediatric Endocrinology | 1 | 0.1% |
| Pediatric Nephrology | 1 | 0.1% |
| Pediatric Surgery | 1 | 0.1% |
| Rheumatology | 1 | 0.1% |
| Rheumatology and Clinical Immunology | 1 | 0.1% |
| Surgical Assists | 1 | 0.1% |
| **Institution / Country of Degree** |  |  |
| UofA - University of Alberta | 158 | 16.5% |
| UofC - University of Calgary | 141 | 14.8% |
| McGill University | 97 | 10.2% |
| University of Ottawa | 70 | 7.3% |
| University of Toronto | 45 | 4.7% |
| University of Saskatchewan | 35 | 3.7% |
| University of British Columbia | 33 | 3.5% |
| Universite de Montreal | 31 | 3.2% |
| Queen's University | 30 | 3.1% |
| Universite de Sherbrooke | 28 | 2.9% |
| McMaster University | 26 | 2.7% |
| Universite Laval | 24 | 2.5% |
| Dalhousie University | 23 | 2.4% |
| University of Manitoba | 23 | 2.4% |
| University of Western Ontario | 18 | 1.9% |
| Egypt | 12 | 1.3% |
| Ireland | 11 | 1.2% |
| Memorial University | 11 | 1.2% |
| Democratic Republic of the Congo | 10 | 1.0% |
| Germany | 9 | 0.9% |
| England | 8 | 0.8% |
| South Africa | 8 | 0.8% |
| Romania | 7 | 0.7% |
| United Kingdom | 7 | 0.7% |
| France | 6 | 0.6% |
| United States of America | 6 | 0.6% |
| Lebanon | 5 | 0.5% |
| Belgium | 4 | 0.4% |
| Iran | 4 | 0.4% |
| The Netherlands | 4 | 0.4% |
| Algeria | 3 | 0.3% |
| Morocco | 3 | 0.3% |
| Netherlands Antilles | 3 | 0.3% |
| Northern Ontario School of Medicine | 3 | 0.3% |
| Poland Poland | 3 | 0.3% |
| Scotland | 3 | 0.3% |
| Spain | 3 | 0.3% |
| Argentina | 2 | 0.2% |
| Australia | 2 | 0.2% |
| Barbados | 2 | 0.2% |
| India | 2 | 0.2% |
| Italy | 2 | 0.2% |
| Mexico Mexico | 2 | 0.2% |
| Physician Lithuania | 2 | 0.2% |
| Poland | 2 | 0.2% |
| Russia | 2 | 0.2% |
| Serbia | 2 | 0.2% |
| Switzerland | 2 | 0.2% |
| Aruba | 1 | 0.1% |
| Chile | 1 | 0.1% |
| Dominican Republic | 1 | 0.1% |
| German Germany | 1 | 0.1% |
| Iceland | 1 | 0.1% |
| Malaysia | 1 | 0.1% |
| Nigeria | 1 | 0.1% |
| Pakistan | 1 | 0.1% |
| Republic of Congo | 1 | 0.1% |
| Saba | 1 | 0.1% |
| Saint Lucia | 1 | 0.1% |
| Saudi Arabia | 1 | 0.1% |
| Sudan | 1 | 0.1% |
| UofC - University of Calgary Undergrads | 1 | 0.1% |
| Wales | 1 | 0.1% |
| West Germany | 1 | 0.1% |
| Yugoslavia | 1 | 0.1% |
| Zaire | 1 | 0.1% |

\* Respondents could report competency in more than one language.

**Appendix D:** Detailed Data Tables, uOttawa Graduates.

| Characteristic (Total n=194) | n | % |
| --- | --- | --- |
| **Gender** |  |  |
| Female | 100 | 51.5% |
| Male | 94 | 48.5% |
| **Language Spoken\*** |  |  |
| English | 194 | 100.0% |
| French | 70 | 36.1% |
| Hindi | 6 | 3.1% |
| Polish | 6 | 3.1% |
| Mandarin | 4 | 2.1% |
| Punjabi | 4 | 2.1% |
| Arabic | 3 | 1.5% |
| Cantonese | 3 | 1.5% |
| Urdu | 3 | 1.5% |
| Bengali | 2 | 1.0% |
| Spanish | 2 | 1.0% |
| Tamil | 2 | 1.0% |
| Vietnamese | 2 | 1.0% |
| Chinese | 1 | 0.5% |
| Cree | 1 | 0.5% |
| German | 1 | 0.5% |
| Hebrew | 1 | 0.5% |
| Hungarian | 1 | 0.5% |
| Italian | 1 | 0.5% |
| Korean | 1 | 0.5% |
| Malayalam | 1 | 0.5% |
| Romanian/Rumanian | 1 | 0.5% |
| Serbo-Croatian | 1 | 0.5% |
| Somali | 1 | 0.5% |
| Ukrainian | 1 | 0.5% |
| **Physician Type** |  |  |
| Community-Based Family Physician | 53 | 27.3% |
| Specialist | 141 | 72.7% |
| **Practice Discipline\*\*** |  |  |
| Family Medicine | 55 | 28.4% |
| Internal Medicine | 30 | 15.5% |
| Anesthesiology | 11 | 5.7% |
| Pediatrics | 11 | 5.7% |
| Diagnostic Radiology | 10 | 5.2% |
| Obstetrics | 10 | 5.2% |
| Emergency Medicine | 7 | 3.6% |
| General Surgery | 7 | 3.6% |
| Ophthalmology | 6 | 3.1% |
| Orthopedic Surgery | 6 | 3.1% |
| Psychiatry | 4 | 2.1% |
| Family Medicine (Emergency Medicine) | 3 | 1.5% |
| Neurology | 3 | 1.5% |
| Anatomical Pathology | 2 | 1.0% |
| Cardiology | 2 | 1.0% |
| Dermatology | 2 | 1.0% |
| Gastroenterology | 2 | 1.0% |
| General Practice | 2 | 1.0% |
| Physical Medicine | 2 | 1.0% |
| Urology | 2 | 1.0% |
| Critical Care Medicine | 1 | 0.5% |
| Family Medicine (Care of the Elderly) | 1 | 0.5% |
| Family Medicine (Sport and Exercise Medicine) | 1 | 0.5% |
| Infectious Diseases | 1 | 0.5% |
| Maternal-Fetal Medicine | 1 | 0.5% |
| Medical Genetics | 1 | 0.5% |
| Medical Oncology | 1 | 0.5% |
| Nephrology | 1 | 0.5% |
| Neurology (Adult and Pediatric) | 1 | 0.5% |
| Neurosurgery | 1 | 0.5% |
| Nuclear Medicine | 1 | 0.5% |
| Otolaryngology - Head and Neck Surgery | 1 | 0.5% |
| Pediatric Neurology | 1 | 0.5% |
| Plastic Surgery | 1 | 0.5% |
| Public Health and Preventive Medicine | 1 | 0.5% |
| Radiation Oncology | 1 | 0.5% |
| Rheumatology | 1 | 0.5% |
| * Respondents could report competency in more than one language. |  |  |
| ** Responses were tabulated as given and may contain inconsistencies. |  |  |

